# The incubation period of COVID-19 – A rapid systematic review and meta-analysis of observational research

**DOI:** 10.1101/2020.04.24.20073957

**Authors:** Conor G. McAloon, Áine B. Collins, Kevin Hunt, Ann Barber, Andrew W. Byrne, Francis Butler, Miriam Casey, John Griffin, Elizabeth Lane, David McEvoy, Patrick Wall, Martin J. Green, Luke O’Grady, Simon J. More

**Affiliations:** Section of Herd Health and Animal Husbandry, UCD School of Veterinary Medicine, University College Dublin, Dublin D04 W6F6, Ireland; Centre for Veterinary Epidemiology and Risk Analysis, UCD School of Veterinary Medicine, University College Dublin, Belfield, Dublin D04 W6F6, Ireland; Centre for Food Safety, UCD School of Biosystems and Food Engineering, University College Dublin, Belfield, Dublin D04 W6F6, Ireland; One Health Scientific Support Unit, Department of Agriculture, Food and the Marine (DAFM), Kildare Street, Dublin 2, Ireland; Woodside Lodge, Barberstown Road, Straffan, County Kildare, Ireland; Department of Agriculture, Food and the Marine, Backweston Campus, Co. Kildare, W23 X3PH, Ireland; School of Public Health, Physiotherapy and Sports Science, Woodview House University College Dublin, Belfield, Dublin D04 W6F6, Ireland; School of Veterinary Medicine and Science, University of Nottingham, Nottingham, UK

**Author notes:** **Correspondence to:** Conor McAloon;, UCD School of Veterinary Medicine, University College Dublin, Dublin, Ireland, 01 716 6083.

**Keywords:** COVID-19, Incubation period, Meta-analysis

## Abstract

**Background:** Reliable estimates of the incubation period are important for decision making around the control of infectious diseases. Knowledge of the incubation period distribution can be used directly to inform decision-making or as inputs into mathematical models.

**Objectives:** The aim of this study was to conduct a rapid systematic review and meta-analysis of estimates of the incubation periods of COVID-19.

**Design:** Rapid systematic review and meta-analysis of observational research

**Data sources:** Publications on the electronic databases PubMed, Google Scholar, MedRxiv and BioRxiv were searched. The search was not limited to peer-reviewed published data, but also included pre-print articles.

**Study appraisal and synthesis methods:** Studies were selected for meta-analysis if they reported either the parameters and confidence intervals of the distributions fit to the data, or sufficient information to facilitate calculation of those values. The majority of studies suitable for inclusion in the final analysis modelled incubation period as a lognormal distribution. We conducted a random effects meta-analysis of the parameters of this distribution.

**Results:** The incubation period distribution may be modelled with a lognormal distribution with pooled mu and sigma parameters of 1.63 (1.51, 1.75) and 0.50 (0.45, 0.55) respectively. The corresponding mean was 5.8 (5.01, 6.69 days). It should be noted that uncertainty increases towards the tail of the distribution: the pooled parameter estimates resulted in a median incubation period of 5.1 (4.5, 5.8) days, whereas the 95^th^ percentile was 11.6 (9.5, 14.2) days.

**Conclusions and implications:** The choice of which parameter values are adopted will depend on how the information is used, the associated risks and the perceived consequences of decisions to be taken. These recommendations will need to be revisited once further relevant information becomes available. Finally, we present an RShiny app that facilitates updating these estimates as new data become available.

**ARTICLE SUMMARY:** *Strengths and limitations of this study:*

- This study provides a pooled estimate of the distribution of incubation periods which may be used in subsequent modelling studies or to inform decision-making
- This estimate will need to be revisited as subsequent data become available. We present an RShiny app to allow the meta-analysis to be updated with new estimates

## INTRODUCTION

Reliable estimates of the incubation period are important for decision making around the control of infectious diseases in human populations. However, incubation periods are expected to vary across individuals within the population. A single measure of central tendency (i.e. mean or median) does not adequately represent this variation accurately.[1] Therefore, it is critically important to understand the variation in incubation periods (i.e. the distribution) within the population.

Knowledge of the incubation period distribution can be used directly to inform decision-making around infectious disease control. For example, the maximum incubation period can be used to inform the duration of isolation, or active monitoring periods of people who have been at high risk of exposure. Knowledge of the incubation period, coupled with estimates of the latent period, serial interval or generation times, may help infer on the duration of the pre-symptomatic infectious period, which is important in understanding both the transmission of infection and opportunities for control.[2] Finally, decision making in the midst of a pandemic often rely on predicted events, such as daily number of new infections, from mathematical models. Such models rely on key input parameters relevant to the transmission of the specific infectious disease. It is important that input parameters into such models are as robust as possible. Given that some models fit data to many parameters, only some of which are specifically of interest but all of which are interdependent, output estimates may be compared to the robust estimates as part of the validation of the model. However, to date, many COVID-19 models have used input values from a single study. The decision on which study to use may vary from model to model. Earlier work has shown that for models of respiratory infections, statements regarding incubation periods are often poorly referenced, inconsistent, or based on limited data.[3]

We hypothesized that a pooled estimate of the distribution of incubation periods could be obtained through a meta-analysis of data published to date. Therefore, the aim of this study was to conduct a rapid systematic review and meta-analysis of estimates of the incubation periods of COVID-19, defined as the period of time (in days) from virus exposure to the onset of symptoms. Specifically, we aimed to find a pooled estimate for the parameters of an appropriate distribution that could be subsequently used as an input in modelling studies and that might help quantify uncertainty around the key percentiles of the distribution as an aid to decision making.

## MATERIALS AND METHODS

For the purpose of this study we followed the Meta-analysis of Observational Studies in Epidemiology (MOOSE) guidelines.[4] The outcome was defined as the time in days from the point of exposure, (in this case, infection) to the onset of clinical signs; all observational studies were included in the analysis. Finally, the population was confirmed infected individuals, where an exposure time could be ascertained with some degree of certainty and precision.

### Search methodology, initial screening and categorisation

A survey of the literature between 1 December 2019 and 8th April 2020 for all countries was implemented using the following search strategy. Publications on the electronic databases PubMed, Google Scholar, MedRxiv and BioRxiv were searched with the following keywords: “Novel coronavirus” OR “SARS□CoV□2” OR “2019-nCoV” OR “COVID-19” AND “incubation period” OR “incubation”. The dynamic curated PubMed database “LitCovid” was also monitored, in addition to national and international government reports. No restrictions on language or publication status were imposed so long as an English abstract was available. Articles were evaluated for data relating to the aim of this review, and all relevant publications were considered for possible inclusion. Bibliographies within these publications were also searched for additional resources. The initial searches were carried out by three of the investigators (ÁC, KH, FB). Authors of studies were contacted only to clarify reporting queries.

### Study appraisal and selection of meta-analysis

Studies were selected for meta-analysis if they reported either the parameters and confidence intervals of the distributions fit to the data, or sufficient information to facilitate calculation of those values. Specifically, this included studies that reported: the point estimate and confidence intervals or standard errors of each parameter; the mean and standard deviation on the original (non-transformed) scale with confidence intervals; the mean and one or more percentiles of the distribution (with confidence intervals); or two or more percentiles of the distribution (with confidence intervals). Studies were excluded if they described the distribution (e.g. with mean, median, percentile) but did not report any uncertainty around that figure. The selection of studies to include in the meta-analysis was conducted by the primary author (CMA).

### Data extraction

On initial appraisal, it was apparent that the majority of studies fitted a lognormal distribution to the data. Earlier work has shown that this distribution is appropriate for many acute infectious diseases.[3, 5] Therefore, the study proceeded as the meta-analysis (pooled estimate) of the parameters of this distribution.

A variable (X) has a lognormal distribution when the log-transformed values follow a normal distribution with mean, mu, and variance, sigma^2, i.e.:

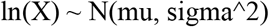

Methods exist for the meta-analysis of studies that combine a mix of log transformed and non-transformed data.[6] In this case we opted to transform data, where possible to the log-transformed scale, and obtain a pooled estimate of both mu and sigma.

### Calculation of distribution parameters from each study

Where the values for each parameter (mu and sigma) were available from the studies, along with corresponding confidence intervals/standard errors, these were extracted as reported. In the remaining studies, the values were calculated where possible from the information presented.

#### Calculation of mu and sigma from studies reporting the mean and standard deviation of the lognormal distribution on the original scale

The mu and sigma parameters of the original lognormal distribution were calculated as:

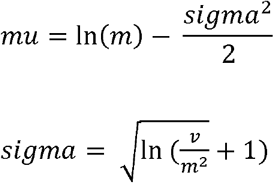

Where *v* = variance (= sd^2^), and *m* = the mean of the distribution on the original (i.e. non-log transformed) scale.

Similarly upper and lower confidence intervals of mu and sigma were found by substituting the upper and lower bounds of the mean or standard deviation (from the original scale) into the equation above, one at a time, whilst holding the value for the other parameter constant (as the point estimate for that parameter).

#### Calculation of mu and sigma from studies reporting mean and percentiles on the original scale

Where studies reported the results as the mean and 95^th^ percentile on the original scale, the “lognorm” package in R was used to calculate the original values of mu and sigma and corresponding standard errors or confidence intervals.[7]

#### Calculation of variance of mu and sigma

For studies reporting confidence intervals, the standard error was calculated as (upper bound – lower bound)/(2 × 1.96)

### Meta-analysis

A random effects meta-analysis was conducted in R-studio Version 1.2.5033,[8] using the “metafor” package,[9] of the mu and sigma parameters of the lognormal distribution, specifying the point estimate and the standard error using “yi” (i.e. the point estimate) and “sei” (i.e. the standard error) arguments. Forest plots were produced using the same package. Quantitative estimates of bias were obtained using the Egger’s test and funnel plots. Heterogeneity was quantified using the I2 statistic and investigated by conducting subgroup analyses of the dataset.

#### Calculation of the se of the mean and sd on the original scale from pooled estimates of mu and sigma

The mean and standard deviation of the pooled estimate were converted to the original (i.e. non-log transformed) scale as:

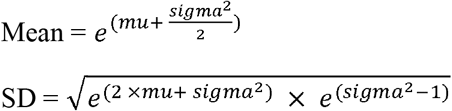

The upper and lower confidence intervals were found by substituting, one at a time, the upper and lower bounds for mu and sigma and recalculating the subsequent figures for mean and SD.

The resulting distribution was plotted using the “ggplot2” package in R.[10] In addition, the distributions for studies that did not fit a lognormal distribution, but that reported the parameters of an alternative distribution fitted were also plotted alongside the pooled lognormal distribution.

Finally, an R Shiny app was created which allows the meta-analysis estimates to be updated as new data become available.

## RESULTS

After initial search and selection of relevant papers and removing duplicates, 20 studies were available for appraisal.

⍰ Two papers were removed as they dealt with specific cohorts of cases – young adults [11] and children.[12]
⍰ One study was removed since only the abstract was in English and there was not enough detail to extract the relevant results.[13]
⍰ Several papers were removed since they contained insufficient data or methods description to facilitate their inclusion:
  - One study was removed since there was not enough detail in the paper to determine whether new parameters were being estimated or whether the parameters quoted were input values for their model.[14]
  - Five papers were removed since the data were largely descriptive, with no confidence intervals reported.[15-19]
  - One study was removed because the error terms associated with the mean, median and percentiles were not reported and there was not enough information presented to recover the parameters of the lognormal distribution.[20]

Of the shortlisted studies (n=10), six reported lognormal distributions as best fitting the data. [21-26] Of the remaining 4, one reported that several distributions were trialled but it was not clear which distribution was used for the final estimates.[27] However, these authors provided raw data which we used to fit the parameters of the lognormal distribution using the “rriskDistributions” package.[28] The remaining 3 studies reported that either Weibull or gamma distributions fitted the data better. Of these, 1 study also presented the results of a log normal distribution fit to the data,[29] facilitating its inclusion in the subsequent analysis. The final two studies reporting a Weibull [30] and a gamma distribution [31] were removed from further analysis at this stage, however, those distributions were plotted over the final distribution to evaluate the impact of removing those studies. The values extracted from each study are shown in Table 1.

**Table 1.**
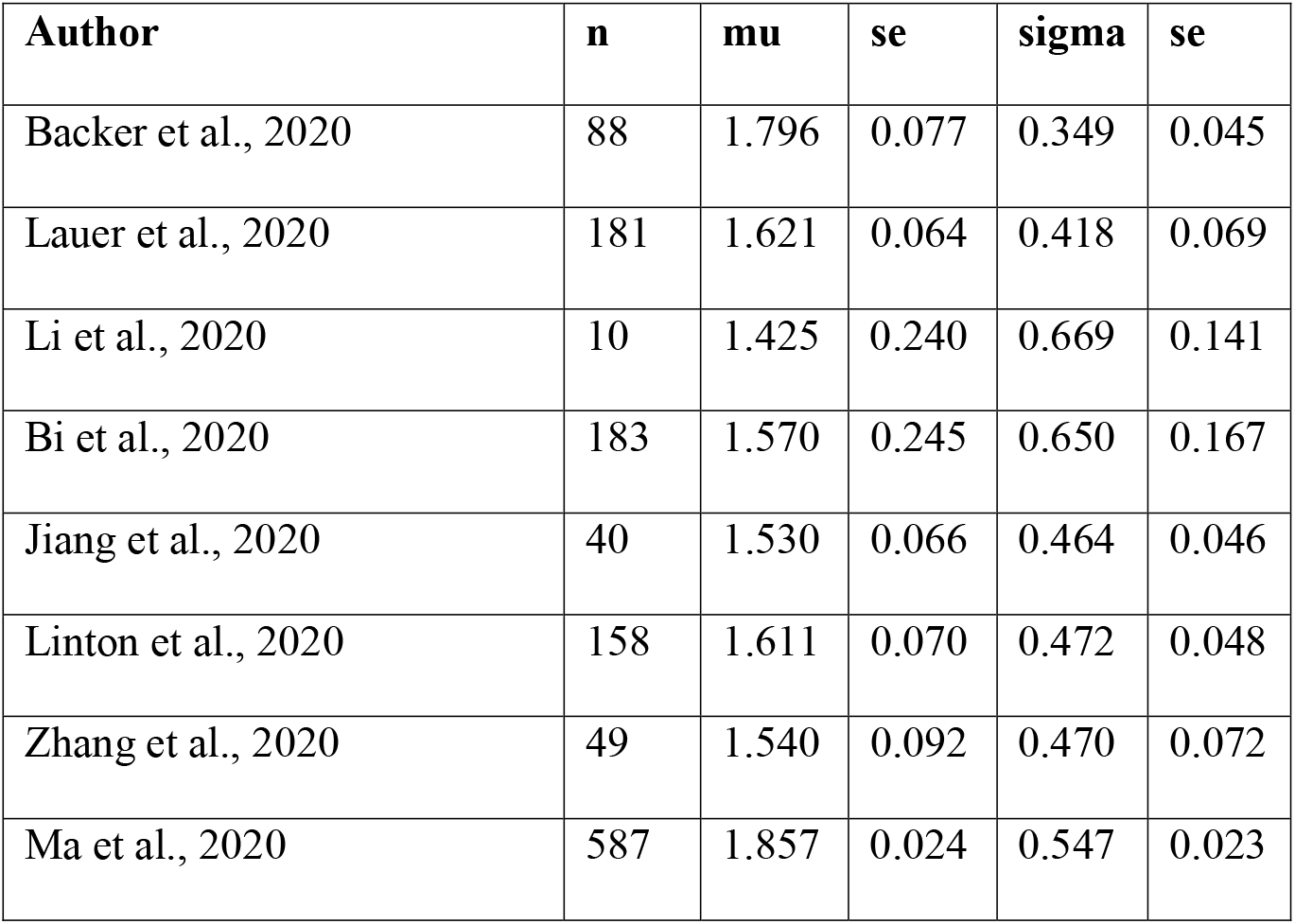
Study size and extracted data for the lognormal mu and sigma parameters from the 8 studies that were used for meta-analysis.

| Author | n | mu | se | sigma | se |
| --- | --- | --- | --- | --- | --- |
| Backer et al., 2020 | 88 | 1.796 | 0.077 | 0.349 | 0.045 |
| Lauer et al., 2020 | 181 | 1.621 | 0.064 | 0.418 | 0.069 |
| Li et al., 2020 | 10 | 1.425 | 0.240 | 0.669 | 0.141 |
| Bi et al., 2020 | 183 | 1.570 | 0.245 | 0.650 | 0.167 |
| Jiang et al., 2020 | 40 | 1.530 | 0.066 | 0.464 | 0.046 |
| Linton et al., 2020 | 158 | 1.611 | 0.070 | 0.472 | 0.048 |
| Zhang et al., 2020 | 49 | 1.540 | 0.092 | 0.470 | 0.072 |
| Ma et al., 2020 | 587 | 1.857 | 0.024 | 0.547 | 0.023 |

The initial pooled estimate of mu from this dataset (i.e. dataset 1, n=8 studies) was 1.65 (1.55, 1.76) and the pooled estimate of sigma was 0.47 (0.41, 0.54). The *I*^*2*^ values were 78% and 59% for mu and sigma respectively. Egger’s tests for mu and sigma were not statistically significant; p=0.11 and p=0.31 for mu and sigma respectively. However, evaluation of the funnel plots (Figures S1 and S2 Supplementary Material) suggests the potential for bias associated with one of the studies included in the analysis.[25] Evaluation of the meta-analyses results for mu demonstrated that two studies were responsible for much of the heterogeneity in the analysis of this value. In particular, the values reported by Ma et al. [25] and Backer et al. [29] were higher than the estimates from other studies. Both studies were further evaluated to determine whether these differences may have been due to methodological differences. The Backer et al. [29] study was subsequently excluded since it appeared that the exposure window was somewhat imprecisely defined which would have biased this estimate upwards. Conversely, the study reported by Ma et al. [25] used only patients where the exposure window was 3 days or less, with the majority of those of a 1-day duration. The meta-analysis was repeated with the Backer et al. [29] study removed (i.e. dataset 2, n=7 studies). The resulting pooled estimates were 1.63 (1.51, 1.75) and 0.50 (0.45, 0.55), whilst the *I*^*2*^ values were 78% and 28% for mu and sigma respectively. Figures 1 and 2 show the resulting forest plots for the meta-analyses of mu and sigma respectively from dataset 2 (n=7), that is the 8 studies from which the parameters were extracted, minus the Backer et al. [29] estimate.

**Figure 1.**
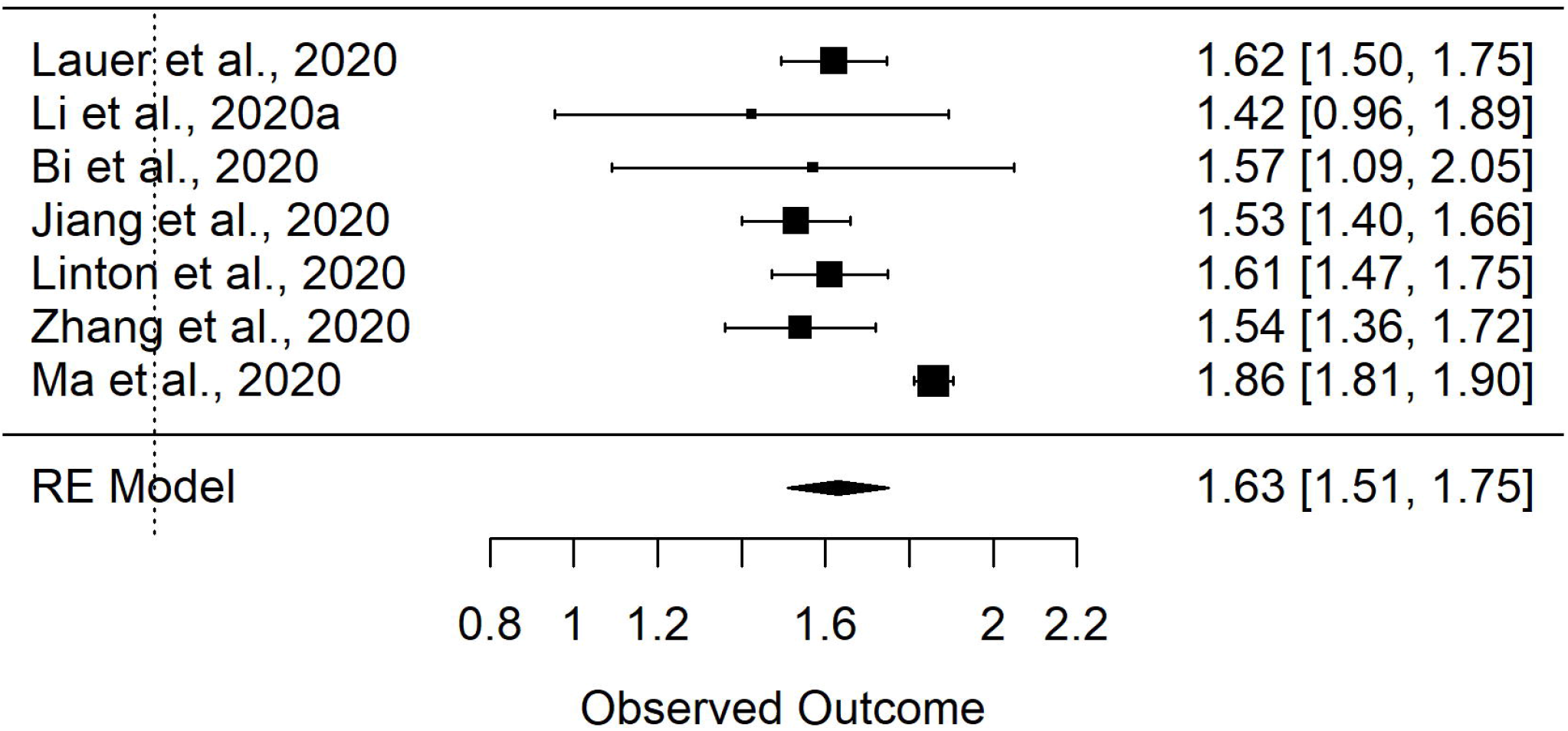
Forest plot of the random effects (RE) meta-analysis of mu parameter of the lognormal distribution of incubation period.

**Figure 2.**
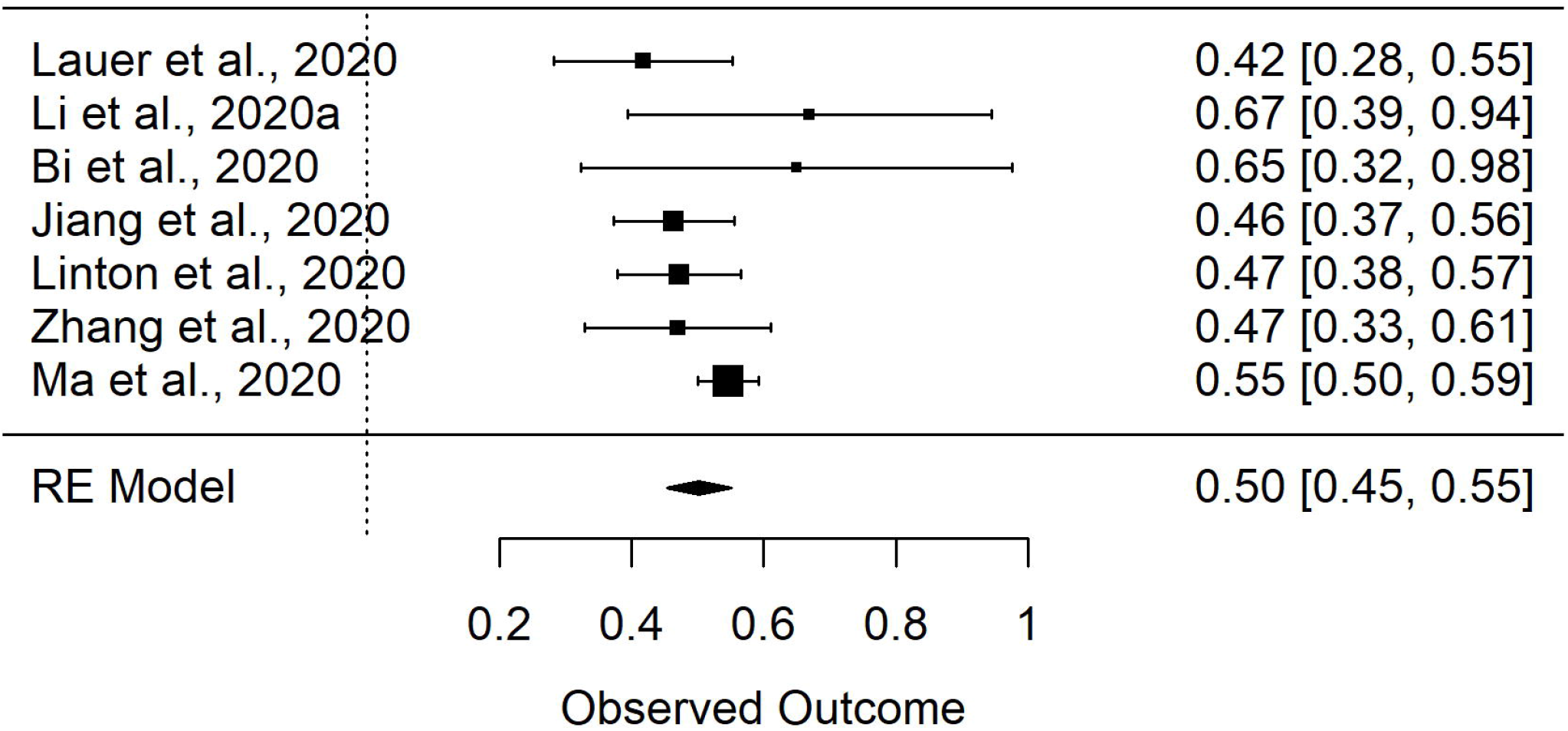
Forest plot of the random effects (RE) meta-analysis of sigma parameter of the lognormal distribution

Figure 3 shows the resulting density plot of the pooled distribution. Figure 4 shows the cumulative density function plot of the same (pooled distribution). In this instance, all possible combinations of distributions across the 95% confidence intervals of the estimates of each of the mu and sigma values are plotted on the same graph. Table 2 shows the percentiles and corresponding confidence intervals of the pooled lognormal distribution.

**Table 2.**
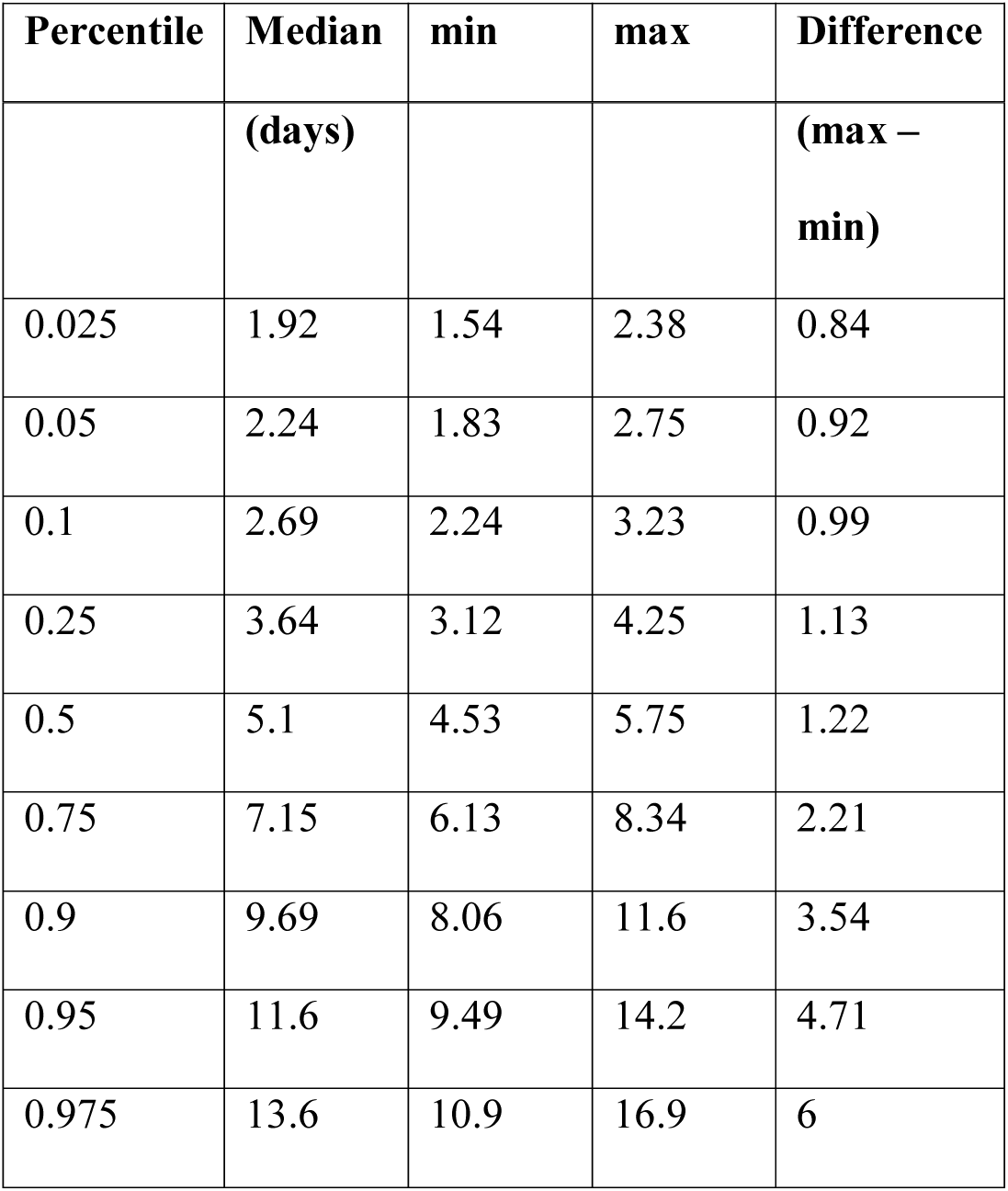
Percentiles of the pooled log normal distribution after simulating all possible combinations of mu and sigma within the 95% confidence intervals of the pooled estimates of both parameters. The median days for each percentile are shown along with the minimum and maximum values for that percentile.

**Figure 3.**
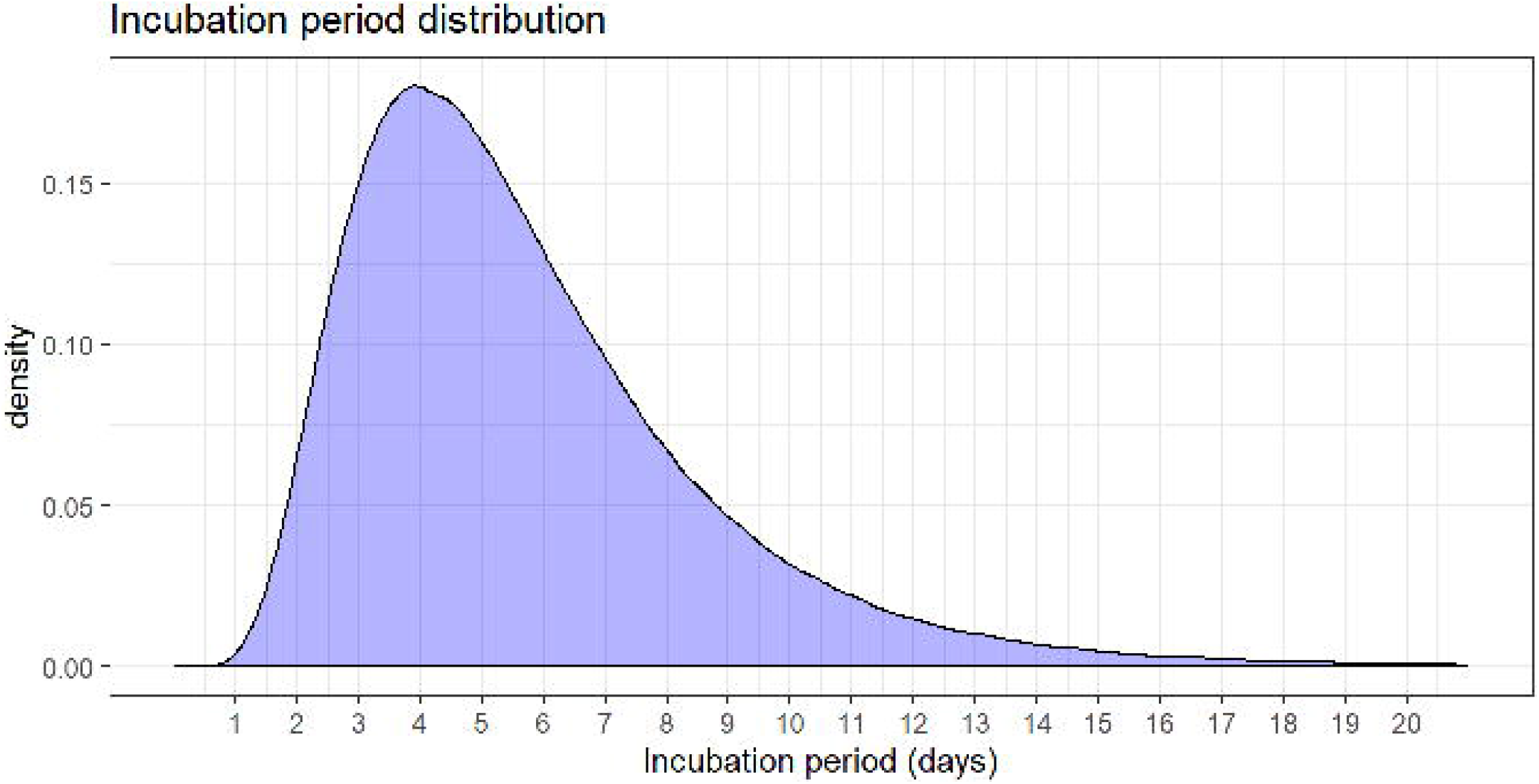
Probability density function of the pooled lognormal distribution of reported incubation period with mu = 1.63 and sigma = 0.50

**Figure 4.**
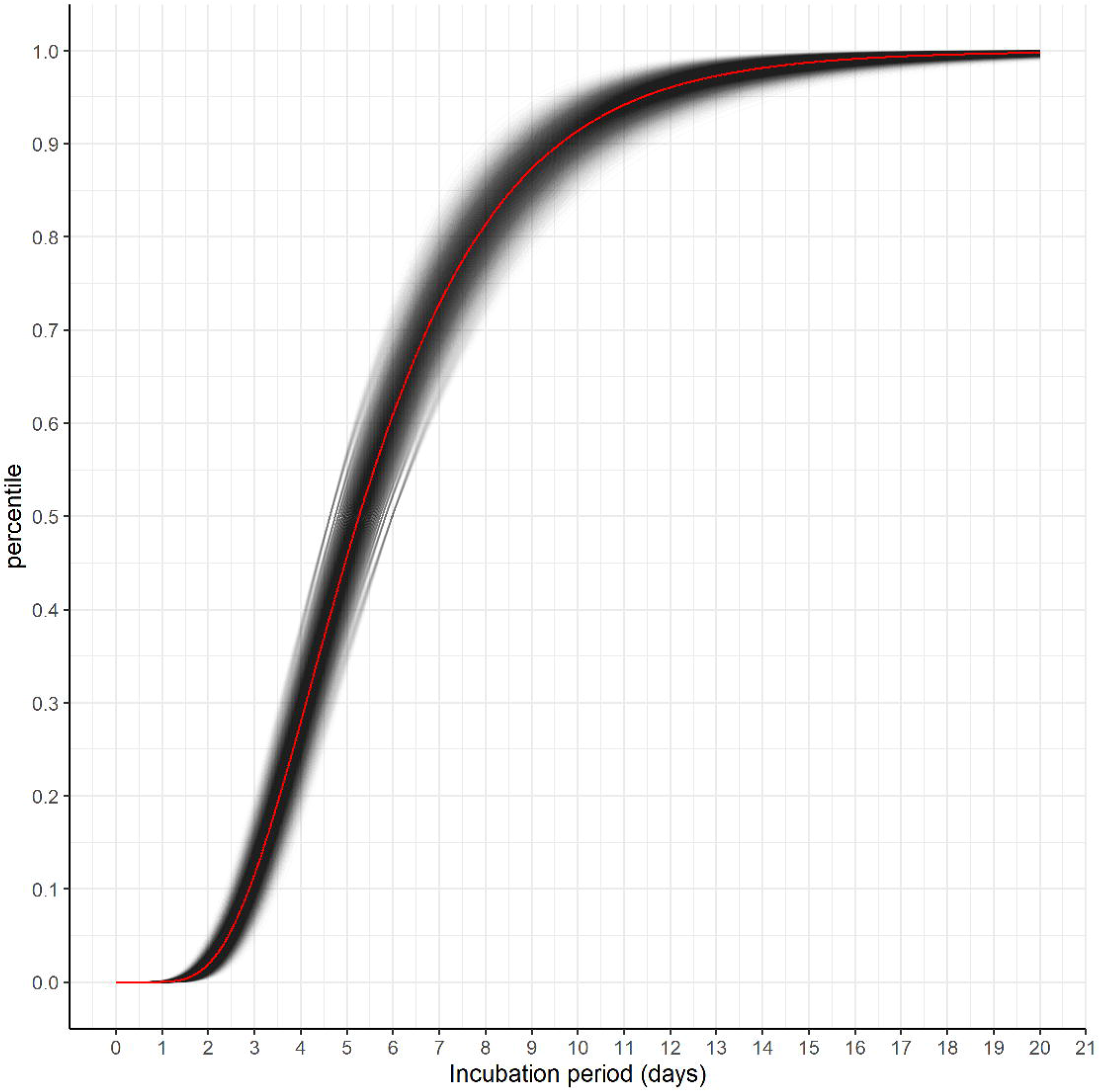
Cumulative distribution function of pooled lognormal distribution. Each possible combination of values between the 95% confidence intervals of mu and sigma are plotted as single black lines.

Figure 5 shows the cumulative density function plots of the pooled lognormal distribution along with the estimates from the original studies. Finally, Figure 6 shows the probability density function of the pooled lognormal distribution, plotted alongside the two studies that could not be included in the final meta-analysis due to the fact that they fit alternative distributions to the data.

**Figure 5.**
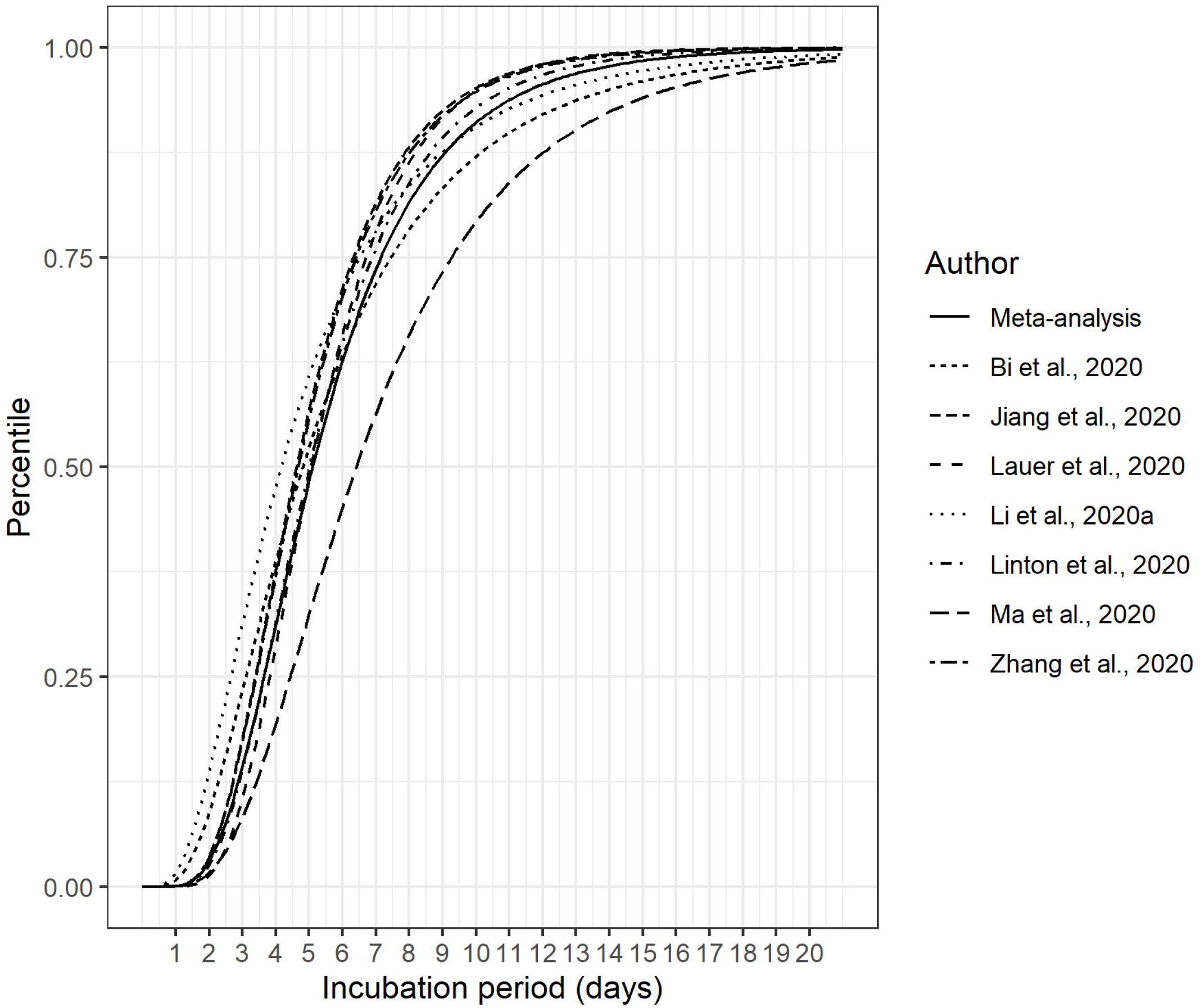
Cumulative distribution function of pooled lognormal distribution for incubation period and original input studies.

**Figure 6.**
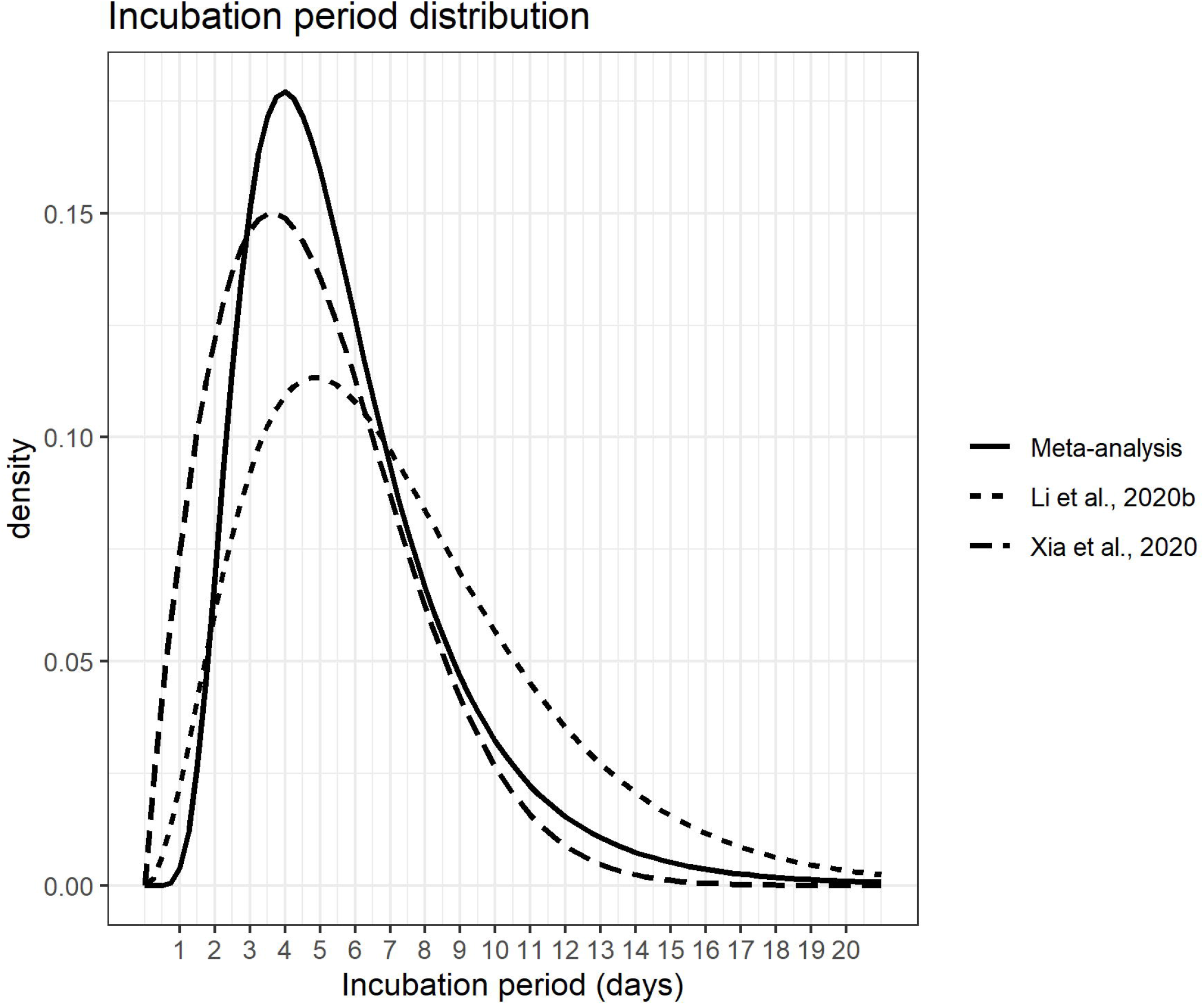
Probability density function of pooled lognormal distribution for incubation period and studies (n=2) not included in the meta-analysis because of the distribution used.

## DISCUSSION

For the purpose of this study we defined incubation period as the time in days from the point of COVID-19 exposure to the onset of symptoms. Figure S1 (Supplementary Material) shows a schematic of this time period with respect to other key parameters influencing COVID-19 transmission. Studies to determine incubation period are likely most precise during the early phase of the outbreak, before the pathogen is widespread.[21] During this early phase, exposure windows can be determined with some confidence. Most studies achieved this by conducting the analysis based on travellers from an epicentre of infection (Wuhan) to another country/region that was free from infection at that time point or in the very early stages of the outbreak.

By definition, the required case data for the determination of individual incubation periods needs to include both exposure (window) and onset of symptoms. Precisely estimating these events can be difficult. Symptom onset is based on case recall, whereas exposure is determined either from: movement history, thereby providing a window prior to movement of potential exposure, or a known window of exposure (from earliest to latest) to a confirmed case (close contact). However, exposure and/or symptom onset are rarely observed exactly. The methods used to deal with this include restricting the analysis to data from patients where the exposure window could be narrowed to a short window (e.g. <3 days); taking a median point from the exposure window to determine the exposure timepoint. Alternatively, Linton et al.[24] included left exposure dates as parameters to be fitted in the model.

After the initial meta-analysis we decided to remove the Backer et al.[29] study from the pooled estimate. The estimates from that study were found to be shifted considerably to the right compared to other estimates. Examination of that study identified that many of the patients had long exposure windows which would be expected to bias the estimate upwards. Interestingly, that study conducted an additional subset analysis of patients whose exposure windows were well defined and for these data, the mean incubation period dropped from 6.4 to 4.5 days. However, it is interesting to note that Ma et al.[25] restricted their analysis to patients with a 3-day exposure window and still found a mean incubation period of 7.4 days. Since this study had the largest sample size (n = 587), it has a significant impact on the estimation of the lognormal parameters. Repeating the meta-analysis with both the Backer et al.[29] and Ma et al.[25] studies removed results in values of 1.58 (1.51, 1.64) and 0.47 (0.42, 0.53) respectively.

With both of these studies removed the *I*^*2*^ values drop to 0% for both parameters. The corresponding mean and median are 5.48 days and 4.85 days respectively. Interestingly, removing this study also increases the precision of the estimate of the value for mu.

One of the weaknesses of our approach is that we extracted and analysed the parameters of the lognormal distribution independently. However, in reality the parameters and the initial distribution that they are fitted to are linked. We were unable to include two studies that did not fit lognormal distributions to the data. However, Figure 6 demonstrates that the impact of removing these studies is likely to be small since they are similar to the pooled estimate, with one falling to the left of the pooled estimate, and the other falling to the right. Ideally, we would have fit distributions to the raw data available from each of the studies, in a way that facilitated the distributions to vary across studies. Such an approach was taken by Lessler et al.[3] in reviewing acute respiratory viral infections. However, the raw data were not available in all cases for the studies that we examined. Another limitation is that many of the papers included in this study used publicly available data to estimate incubation period. Therefore, there is a reasonable chance that several of the analyses have re-used at least some of the same data. In these cases, the studies would not be independent of each other.

It is worth noting that the parameter values from our meta-analysis are somewhat higher than previously used in modelling studies. For example, Ferguson et al.[32] used a mean of 5.1 days for incubation period, citing two previous studies.[24, 31] Mean incubation period from our meta analysis was 5.8. Tuite et al.[33] on the other hand, used an incubation period of 5.0 days citing the study by Lauer et al.[22]. This figure, (5.0 days) was the median incubation period reported from that study,[22] which is much closer to the median estimate of 5.1 days from our meta analysis.

It is reasonable to assume that the incubation period estimated here should be relatively generalizable across different populations: unlike parameters such as serial interval for example, incubation period depends only on the interaction between the virus and the host, which is expected to be similar across populations, and not on behavioural factors such as frequency of contacts which might be expected to vary across different countries. However, there is potential for a number of biases in these data which may impact on their external validity: In order to accurately estimate incubation period, it is possible that well characterized cases which may be preferentially chosen to reduce the impact of prolonged exposure windows. It is possible that such cases could be biased towards more severe cases. In that case, the estimate for incubation period could be biased downwards, since it is possible that the incubation period could be shorter in more severely affected individuals. Furthermore, these well characterised cases may not have been representative of all cases (often male, often younger,[29]), highlighting the need for information on incubation period from older people, people with comorbidities, from women and those with mild symptoms. These findings are mostly based on studies from Chinese patients. Whilst the incubation period for a given set of circumstances should be similar across different populations, there may be factors that might impact on incubation period, such as infectious dose for example that might vary between populations (and possibly within populations over the course of the outbreak) meaning that the resulting distribution may vary for different populations, or potentially at different stages of the outbreak. Finally, incubation periods may be different for people of different ages.[11]

Based on available evidence, we find that the incubation period distribution may be modelled with a lognormal distribution with pooled mu and sigma parameters of 1.63 (1.51, 1.75) and 0.50 (0.45, 0.55) respectively. It should be noted that uncertainty increases towards the tail of the distribution (Figure 4 and Table 2). The choice of which parameter values are adopted will depend on how the information is used, the associated risks and the perceived consequences of decisions to be taken. The corresponding mean was 5.8 days and the median was 5.1 days. These recommendations will need to be revisited once further relevant information becomes available. Finally, we present an R Shiny app which facilitates users to update these estimates as new data become available https://mcaloon-ucd.shinyapps.io/shiny2/.

## Data Availability

The data for these meta-analyses are presented as part of the manuscript

## Author contributions

CMA conducted the eligibility screening of shortlisted studies, extracted the data and conducted the analysis with input from all authors; ÁC, KH and FB conducted the initial literature searches; CMA and SM completed the initial drafts of the manuscript; MG and LOG reviewed the statistical methods; All authors read and approved the final manuscript.

## Data statement

The data for the meta-analyses are presented as part of the manuscript (Table 2).

## Patient and public involvement statement

It was not appropriate or possible to involve patients or the public in the design, or conduct, or reporting, or dissemination plans of our research

